# Community-acquired pneumonia identification from electronic health records in the absence of a gold standard: a Bayesian latent class analysis

**DOI:** 10.1101/2025.02.03.25321566

**Authors:** Jia Wei, Kevin Yuan, Augustine Luk, A Sarah Walker, David W Eyre

**Affiliations:** Nuffield Department of Medicine, University of Oxford, Oxford, UK; Big Data Institute, Nuffield Department of Population Health, University of Oxford, Oxford, UK; The National Institute for Health Research Oxford Biomedical Research Centre, University of Oxford, Oxford, UK; The National Institute for Health Research Health Protection Research Unit in Healthcare Associated Infections and Antimicrobial Resistance at the University of Oxford, Oxford, UK; Department of Infectious Diseases and Microbiology, Oxford University Hospitals NHS Foundation Trust, John Radcliffe Hospital, Oxford, UK

## Abstract

**Background:** Community-acquired pneumonia (CAP) is common and a significant cause of mortality. However, CAP surveillance commonly relies on diagnostic codes from electronic health records (EHRs), whose accuracy is imperfect.

**Methods:** We used Bayesian latent class models to assess the accuracy of CAP diagnostic codes in the absence of a gold standard and to explore the contribution of various EHR data sources in improving CAP identification. Using records from 491,681 hospital admissions in Oxfordshire, UK, from 2016 to 2023, we investigated four EHR-based algorithms for CAP detection based on 1) primary diagnostic codes, 2) clinician-documented indications for antibiotic prescriptions, 3) radiology free-text reports, and 4) vital signs and blood tests.

**Results:** The estimated prevalence of CAP as the reason for emergency hospital admission was 13.2% (95% credible interval 12.8-13.6%). Primary diagnostic codes had low sensitivity but a high specificity (best fitting model, 0.283 and 0.997 respectively), as did vital signs with blood tests (0.242 and 0.988). Antibiotic indication text had a higher sensitivity (0.603) but a lower specificity (0.981), with radiology reports intermediate (0.493 and 0.959). Defining CAP as present when detected by any algorithm produced sensitivity and specificity of 0.854 and 0.925 respectively. Results remained consistent using alternative priors and in sensitivity analyses.

**Conclusion:** Relying solely on diagnostic codes for CAP surveillance leads to substantial under-detection; combining EHR data across multiple algorithms enhances identification accuracy. Bayesian latent class analysis-based approaches could improve CAP surveillance and epidemiological estimates by integrating multiple EHR sources, even without a gold standard for CAP diagnosis.

## Introduction

Community-acquired pneumonia (CAP) is a leading cause of death globally^1^, and a major driver of antimicrobial use and resistance^2^. Identifying CAP in epidemiological and clinical research often relies on diagnostic codes from electronic health records (EHRs)^3^, however, accuracy remains in question. Diagnostic codes are primarily recorded for administrative purposes, e.g. hospital performance monitoring and insurance reimbursement, rather than epidemiology; this will inevitably influence a hospital’s coding practice. Furthermore, due to clinical uncertainty or multiple possible diagnoses, the choice of diagnostic codes is also influenced by coders’ experience and documentation quality^4^. Previous studies have reported low accuracy of using administrative data for CAP identification, with sensitivity ranging from 0.38 to 0.75^5–12^. Thus, relying solely on diagnostic codes can introduce bias in some research contexts^13^. Examining the accuracy of diagnostic codes and validating their use before performing epidemiological analyses is warranted^14^, alongside exploring alternative strategies for identifying CAP.

The traditional method for validating a diagnostic test or code involves comparing it against a gold standard with assumed perfect accuracy. However, CAP definitions and diagnostic criteria vary significantly across different healthcare settings and countries, making a universally accepted gold standard difficult to establish^15,16^. This partly reflects the challenging nature of identifying CAP for clinicians, especially in patients presenting with non-specific symptoms and signs, and with non-diagnostic imaging. This can lead to imprecise surveillance estimates, poorly classified clinical phenotypes, and inaccurate clinical decision making, leading to inappropriate antibiotic use, sub-optimal patient outcomes, and subsequent antibiotic resistance^17^. Therefore, improving the accuracy of CAP identification is essential for effective management, disease surveillance, and combating antimicrobial resistance.

In this study, we applied Bayesian latent class analysis, a method that allows for diagnostic validation in the absence of a gold standard, to assess the accuracy of CAP diagnostic codes. We also examined if other routinely collected EHR data could improve CAP identification.

## Methods

### Population

We used de-identified electronic patient record data from Oxford University Hospitals (OUH) NHS Foundation Trust available within the Infections in Oxfordshire Research Database (IORD). IORD has Research Ethics Committee, Health Research Authority and Confidentiality Advisory Group approvals (19/SC/0403,19/CAG/0144) as a de-identified research database without individual consent. We included all adults (≥16 years) who entered the hospital through the Emergency Department or other emergency admissions units (determined by NHS admission method code^18^) from January 2016 to December 2023 inclusive. Admissions are recorded as one or more episodes during which a different senior doctor or specialty is responsible for care. We only included the first episode among each admission to focus on CAP, i.e. only considering patients where pneumonia was potentially the reason for hospital admission. Admissions with missing patient identifiers or missing diagnostic codes were excluded, as were admissions with a SARS-CoV-2 infection diagnosis code (U07.1/U07.2) and from 01-February-2020 to 31-May-2020 (i.e., prior to widespread SARS-CoV-2 testing) to avoid including patients with COVID-19 pneumonia (**Figure 1**).

**Figure 1.**
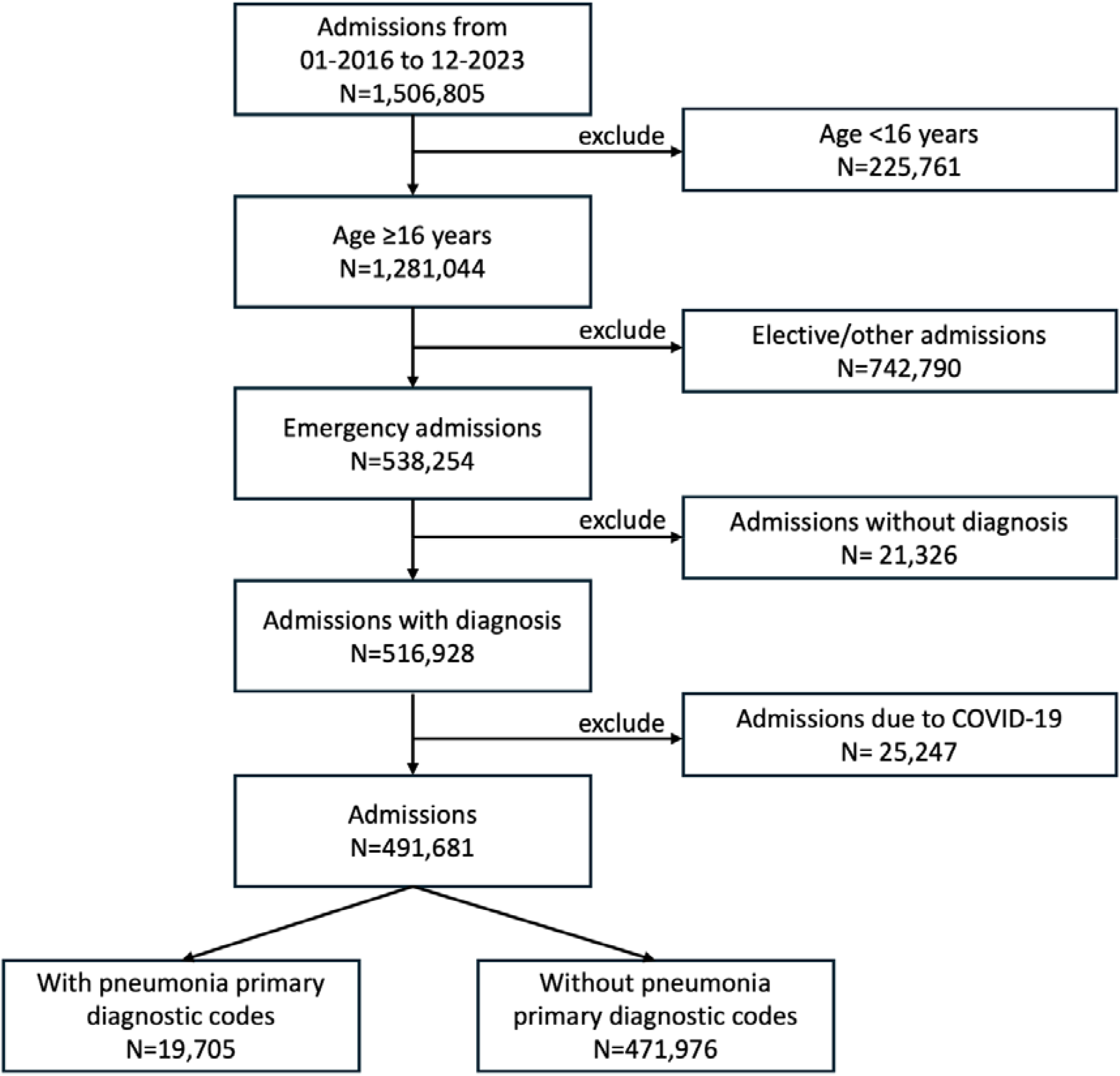
Study inclusion and exclusion flowchart.

### Algorithms for identifying CAP

We investigated four algorithms using EHR data to identify CAP cases.

#### Primary diagnostic codes (algorithm-1, codes)

We extracted the primary diagnostic codes from the first episode of each admission (International Classification of Diseases, Tenth Revision, Clinical Modification, ICD-10-CM) and defined CAP by any of: J189, J181, J180, J159, J13, J151, J154, J851, J14, J152, A481, J150, J188, J157, J155, J156, J850, J158, J153, J168. J189 (Pneumonia, unspecified organism) and J181 (Lobar pneumonia, unspecified organism) accounted for 96% of cases (**Table S1**).

#### Indication in antibiotic prescriptions (algorithm-2, indication)

EHR antibiotic prescriptions in our dataset include a free-text field indicating the reason for prescribing. We used a previously reported transformer model to identify sources of infection from this text^19^. Then, among those with a respiratory source identified, we used fuzzy string-matching to find indications consistent with CAP, i.e., if any of the following search terms appeared in the free-text: “pneumonia”, “CAP”, “consolidation”, “LRTI (lower respiratory tract infection)”, “chest”, and “CURB”. All antibiotic prescriptions within [-2, +48] hours of linked admission times were used.

#### Free text radiology reports (algorithm-3, radiology)

The first chest X-ray (CXR) or computed tomography (CT) report within [-48,+48] hours of each admission time was selected. Weak labels, i.e., designed to screen permissively for presence/absence of CAP, were assigned using the following labelling rules (given in detail in **Supplementary Methods**). Admissions without radiology scans within the timeframe above were assumed to indicate absence of CAP in the primary analysis. In sensitivity analysis, admissions without radiology scans were excluded from analysis.

#### Shortness of breath and elevated CRP (algorithm-4, tests)

CAP patients often present with shortness of breath and elevated C-reactive protein (CRP) levels, with previous studies demonstrating CRP’s diagnostic value for CAP^20–22^. Therefore, we heuristically defined CAP by having shortness of breath (evidenced by oxygen saturation <90% or respiratory rate >25 breaths per minute or requirement for supplemental inspired oxygen) and an elevated CRP level (>5 mg/L) using the first vital signs and laboratory measurements within [-2,+24] hours of admission time. In the primary analysis, admissions with missing vital signs and CRP measurements were assumed to indicate absence of CAP. In sensitivity analyses, missing vital signs and CRP measurements were imputed using multiple imputation by chained equations (MICE) or excluded from analysis.

### Statistical analysis

Without a perfect reference standard, the true disease status is ‘latent’, meaning it cannot be directly observed. A Bayesian latent class model (BLCM) can estimate the accuracy of diagnostic tests without relying on information from a reference test^23^. The performance of each diagnostic test is represented by a set of parameters, with the uncertainty around performance represented by probability distributions around each parameter value. Prior beliefs about each diagnostic test’s performance (parameter priors) (details in **Table S3**) are combined with the likelihood of obtaining the observed data at each possible set of parameter values to obtain a posterior distribution on the likely values for each parameter (summarized with point estimates and credibility intervals), using a principled search of all possible parameter values and latent states. A multinomial distribution was assumed for the cross-classified results of the four algorithms, a commonly used method to model cross-classified binary diagnostic tests^24,25^.

We initially constructed a BLCM assuming conditional independence among the four algorithms (Model-1). However, as results from different tests may be correlated, additional models incorporated fixed-effect conditional dependence^26^. For Model-2, we added pairwise conditional dependence between Algorithms-1 (codes) and -2 (indication), as well as Algorithms-1 (codes) and -3 (radiology). For Model-3, we further added pairwise conditional dependence between Algorithm-1 (codes) and -4 (tests) in CAP patients. For Model-4, we further assumed that diagnostic codes and antibiotic indication were also correlated among non-CAP patients.

We calculated the sensitivity, specificity, positive predictive value (PPV), and negative predictive value (NPV) of each algorithm. We used the Deviance information criterion (DIC), an estimate of expected predictive error, to compare models and identify those with the best fit^27^. Details of prior choices and computation are provided in **Supplementary Methods**.

## Results

Among 491,681 admissions (**Figure 1**), 19,705 (4.0%) had a primary CAP diagnostic code, 47,301 (9.6%) had an antibiotic prescription indication consistent with CAP, 49,445 (10.1%) had CAP identified in a CXR or CT report (276,219 (56.2%) had no scan performed, assumed no CAP in primary analysis) and 20878 (4.2%) patients presented with shortness of breath and elevated CRP levels (110,884 (22.6%) had missing values for either, assumed normal in primary analysis).

Of the four models fitted (see Methods), Models-2, -3, and -4, which included correlations between the different approaches, all had lower DIC (i.e. better model fit) than Model-1 which assumed conditional independence (**Table 1**). Gelman-Rubin statistics and MCMC trace plots showed good convergence for all models (**Figure S1**). Model-3 had the lowest DIC (246) and was simpler than Model-4, so was used for all analyses.

**Table 1.**
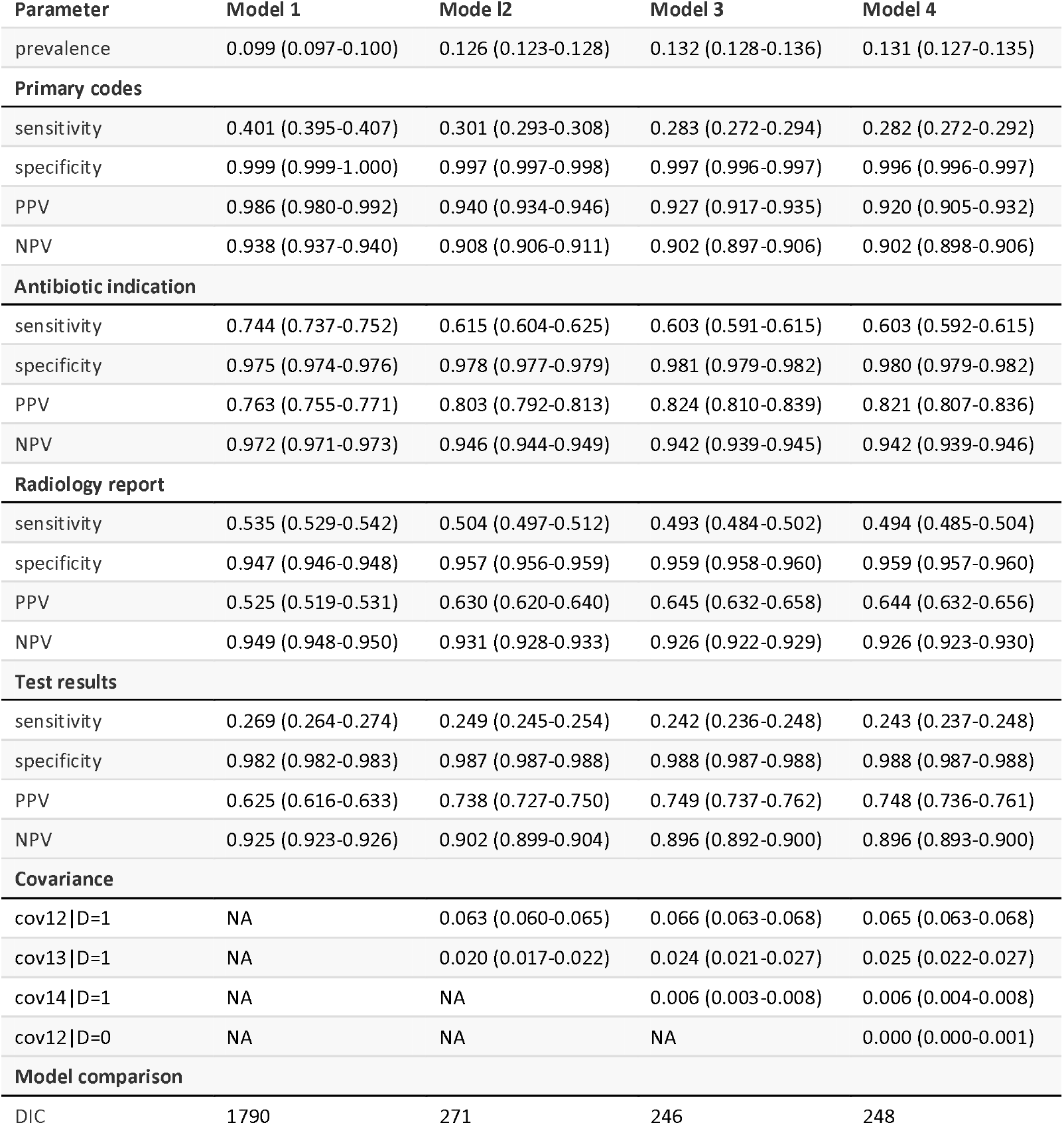
Posterior predicted sensitivity, specificity, PPV, NPV, covariance, and model comparison among four models with different covariance structures. PPV: positive predictive value; NPV: negative predictive value; DIC: deviance information criteria.

| Parameter | Model 1 | Model 2 | Model 3 | Model 4 |
| --- | --- | --- | --- | --- |
| prevalence | 0.099 (0.097-0.100) | 0.126 (0.123-0.128) | 0.132 (0.128-0.136) | 0.131 (0.127-0.135) |
| **Primary codes** |  |  |  |  |
| sensitivity | 0.401 (0.395-0.407) | 0.301 (0.293-0.308) | 0.283 (0.272-0.294) | 0.282 (0.272-0.292) |
| specificity | 0.999 (0.999-1.000) | 0.997 (0.997-0.998) | 0.997 (0.996-0.997) | 0.996 (0.996-0.997) |
| PPV | 0.986 (0.980-0.992) | 0.940 (0.934-0.946) | 0.927 (0.917-0.935) | 0.920 (0.905-0.932) |
| NPV | 0.938 (0.937-0.940) | 0.908 (0.906-0.911) | 0.902 (0.897-0.906) | 0.902 (0.898-0.906) |
| **Antibiotic indication** |  |  |  |  |
| sensitivity | 0.744 (0.737-0.752) | 0.615 (0.604-0.625) | 0.603 (0.591-0.615) | 0.603 (0.592-0.615) |
| specificity | 0.975 (0.974-0.976) | 0.978 (0.977-0.979) | 0.981 (0.979-0.982) | 0.980 (0.979-0.982) |
| PPV | 0.763 (0.755-0.771) | 0.803 (0.792-0.813) | 0.824 (0.810-0.839) | 0.821 (0.807-0.836) |
| NPV | 0.972 (0.971-0.973) | 0.946 (0.944-0.949) | 0.942 (0.939-0.945) | 0.942 (0.939-0.946) |
| **Radiology report** |  |  |  |  |
| sensitivity | 0.535 (0.529-0.542) | 0.504 (0.497-0.512) | 0.493 (0.484-0.502) | 0.494 (0.485-0.504) |
| specificity | 0.947 (0.946-0.948) | 0.957 (0.956-0.959) | 0.959 (0.958-0.960) | 0.959 (0.957-0.960) |
| PPV | 0.525 (0.519-0.531) | 0.630 (0.620-0.640) | 0.645 (0.632-0.658) | 0.644 (0.632-0.656) |
| NPV | 0.949 (0.948-0.950) | 0.931 (0.928-0.933) | 0.926 (0.922-0.929) | 0.926 (0.923-0.930) |
| **Test results** |  |  |  |  |
| sensitivity | 0.269 (0.264-0.274) | 0.249 (0.245-0.254) | 0.242 (0.236-0.248) | 0.243 (0.237-0.248) |
| specificity | 0.982 (0.982-0.983) | 0.987 (0.987-0.988) | 0.988 (0.987-0.988) | 0.988 (0.987-0.988) |
| PPV | 0.625 (0.616-0.633) | 0.738 (0.727-0.750) | 0.749 (0.737-0.762) | 0.748 (0.736-0.761) |
| NPV | 0.925 (0.923-0.926) | 0.902 (0.899-0.904) | 0.896 (0.892-0.900) | 0.896 (0.893-0.900) |
| **Covariance** |  |  |  |  |
| cov12 \| D=1 | NA | 0.063 (0.060-0.065) | 0.066 (0.063-0.068) | 0.065 (0.063-0.068) |
| cov13 \| D=1 | NA | 0.020 (0.017-0.022) | 0.024 (0.021-0.027) | 0.025 (0.022-0.027) |
| cov14 \| D=1 | NA | NA | 0.006 (0.003-0.008) | 0.006 (0.004-0.008) |
| cov12 \| D=0 | NA | NA | NA | 0.000 (0.000-0.001) |
| **Model comparison** |  |  |  |  |
| DIC | 1790 | 271 | 246 | 248 |

The estimated prevalence of CAP as the reason for emergency admission to hospital was 13.2% (95% credible interval [CrI] 12.8-13.6%). The estimated sensitivity of primary diagnostic codes was only 0.283 (95% CrI 0.272-0.294), but the specificity was very high at 0.997 (95% Crl 0.996-0.997). Antibiotic indications had a higher sensitivity of 0.603 (95% CrI 0.591-0.615) with a specificity of 0.981 (0.979-0.982). The sensitivity and specificity of radiology was 0.493 (95% CrI 0.484-0.502), and 0.959 (95% CrI 0.958-0.960), respectively. The algorithm based on tests (shortness of breath and elevated CRP) had the lowest sensitivity of 0.242 (95% CrI: 0.236-0.248), but relatively high specificity at 0.988 (95% CrI: 0.987-0.988). Although all four algorithms demonstrated high specificity, sensitivity was generally lower, with the trade-off of slightly reduced specificity for algorithms with higher sensitivity. The negative predictive value (NPV) for all algorithms was similar, with the antibiotic indication algorithm achieving the highest NPV at 0.942 (95% CrI: 0.939-0.945). Primary diagnostic codes had the highest positive predictive value (PPV) at 0.927 (95% CrI: 0.917-0.935) (**Figure 2a, b, Table 1**).

**Figure 2.**
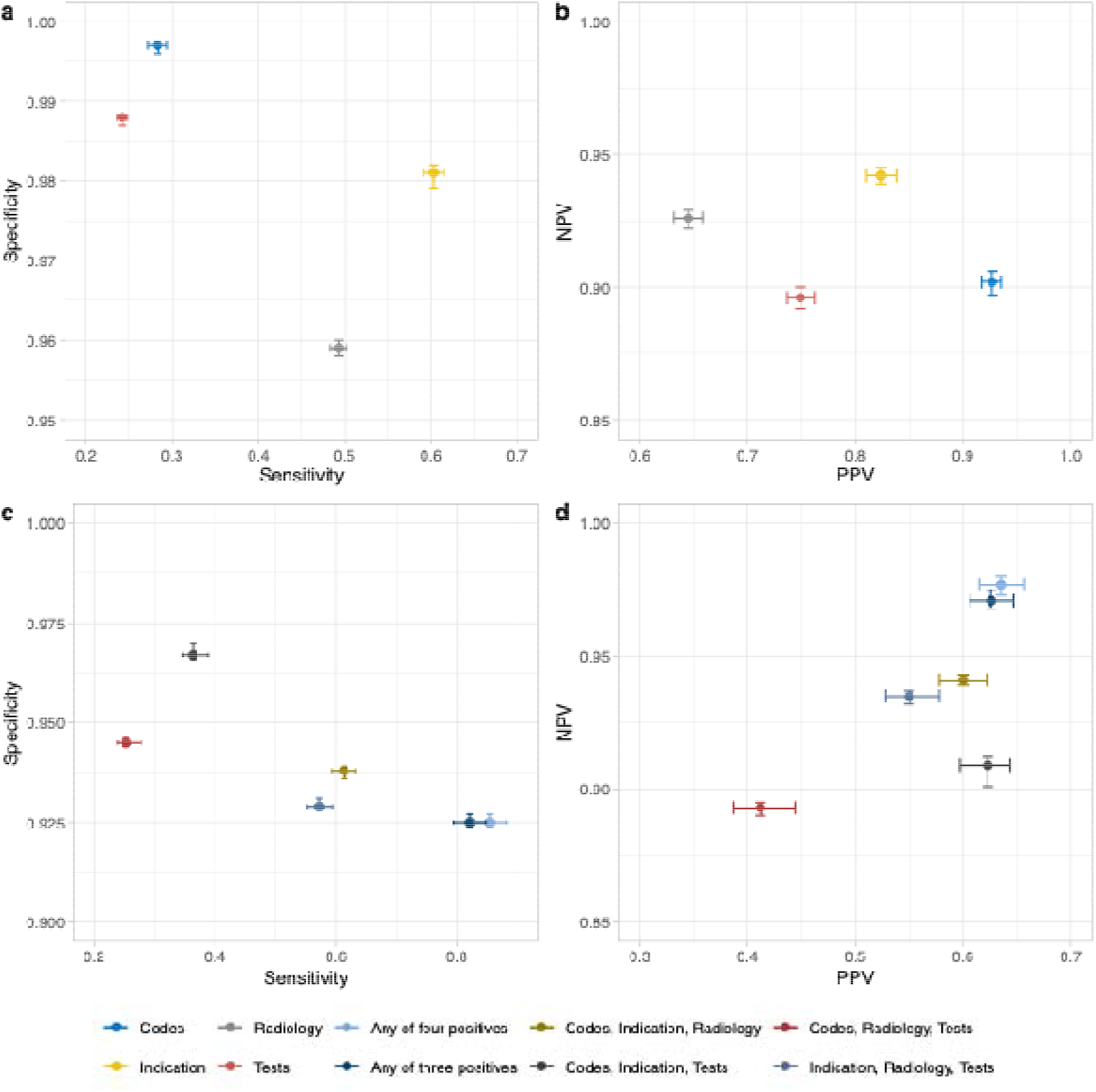
Predicted sensitivity, specificity, PPV, and NPV of the four algorithms (a, b) and algorithms combinations (c, d) under Model-3 in the primary analysis. Algorithm combinations used an ‘OR’ rule for three or four positive outcomes. PPV: positive predictive value; NPV: negative predictive value. Codes: CAP primary diagnostic codes; Indication: CAP antibiotic indication; Radiology: chest X-ray report or CT scan report; Tests: shortness of breath and elevated C-reactive protein levels.

Among 11 combinations of approaches for detecting pneumonia (two, three, or four positives), the most common was antibiotic indication and radiology (N=8,559), followed by diagnostic code, antibiotic indication, and radiology (N=6,419). Predicted frequency of each combination under Model-3 aligned well with the observed frequency (**Figure 3**). Results were generally consistent across other Models-1, -2, and -4 (**Table S4**). Using an ‘AND’ rule that required at least 2 approaches to detect pneumonia, all combinations of approaches had low sensitivity (0.009-0.127) but high specificity (all close to 1), high PPV (0.922-1.00) and somewhat lower NPV (0.869-0.888). The combined sensitivity and specificity of requiring any three positives was 0.172 and 1.000, and of requiring any two positives was 0.318 and 1.000 (**Table 2, Figure S2)**. Using an ‘OR’ rule, i.e. requiring just one of several approaches, the sensitivity of the outcome combinations was higher, but as expected the PPV was lower than the ‘AND’ rule. For example, the sensitivity, specificity, PPV, and NPV of requiring any one of the four approaches were 0.854, 0.925, 0.635, and 0.977 (**Figure 2c, d, Table 2**).

**Table 2.** Posterior predicted sensitivity, specificity, PPV, NPV for each algorithm combination under the ‘AND’ rule and ‘OR’ rule in Model-3 in the primary analysis. PPV: positive predictive value; NPV: negative predictive value. Numbers are presented by upset plot in **Figure S2**.

| Combinations | 'AND' rule |  |  |  | 'OR' rule |  |  |  |
| --- | --- | --- | --- | --- | --- | --- | --- | --- |
|  | Sensitivity | Specificity | PPV | NPV | Sensitivity | Specificity | PPV | NPV |
| **Codes, Indication, Radiology, Tests**<br>(‘AND’ requires all 4 be positive,<br>‘OR’ requires ≥1 be positive) | 0.033 | 1.000 | 1.000 | 0.872 | 0.854 | 0.925 | 0.635 | 0.977 |
| **Codes, Indication, Radiology** | 0.097 | 1.000 | 1.000 | 0.879 | 0.613 | 0.938 | 0.600 | 0.941 |
| **Codes, Indication, Tests** | 0.027 | 1.000 | 1.000 | 0.871 | 0.363 | 0.967 | 0.623 | 0.909 |
| **Codes, Radiology, Tests** | 0.009 | 1.000 | 0.999 | 0.869 | 0.253 | 0.945 | 0.412 | 0.893 |
| **Indication, Radiology, Tests** | 0.038 | 1.000 | 0.998 | 0.872 | 0.572 | 0.929 | 0.550 | 0.935 |
| **Any three positives**<br>(‘AND’ requires any 3 be positive,<br>‘OR’ requires 1 of the 4 be negative) | 0.172 | 1.000 | 1.000 | 0.888 | 0.821 | 0.925 | 0.626 | 0.971 |
| **Codes, Indication** | 0.078 | 1.000 | 0.995 | 0.877 | 0.240 | 0.978 | 0.628 | 0.894 |
| **Codes, Radiology** | 0.023 | 1.000 | 0.963 | 0.871 | 0.158 | 0.957 | 0.358 | 0.882 |
| **Indication, Radiology** | 0.127 | 0.999 | 0.961 | 0.883 | 0.405 | 0.941 | 0.511 | 0.912 |
| **Codes, Tests** | 0.005 | 1.000 | 0.946 | 0.869 | 0.058 | 0.985 | 0.375 | 0.873 |
| **Indication, Tests** | 0.047 | 1.000 | 0.969 | 0.873 | 0.244 | 0.970 | 0.551 | 0.894 |
| **Radiology, Tests** | 0.038 | 1.000 | 0.922 | 0.872 | 0.207 | 0.948 | 0.378 | 0.887 |
| **Any two positives**<br>(‘AND’ required any 2 be positive,<br>‘OR’ requires 2 of the 4 be negative) | 0.318 | 0.998 | 0.965 | 0.906 | 0.649 | 0.925 | 0.570 | 0.945 |

**Figure 3.**
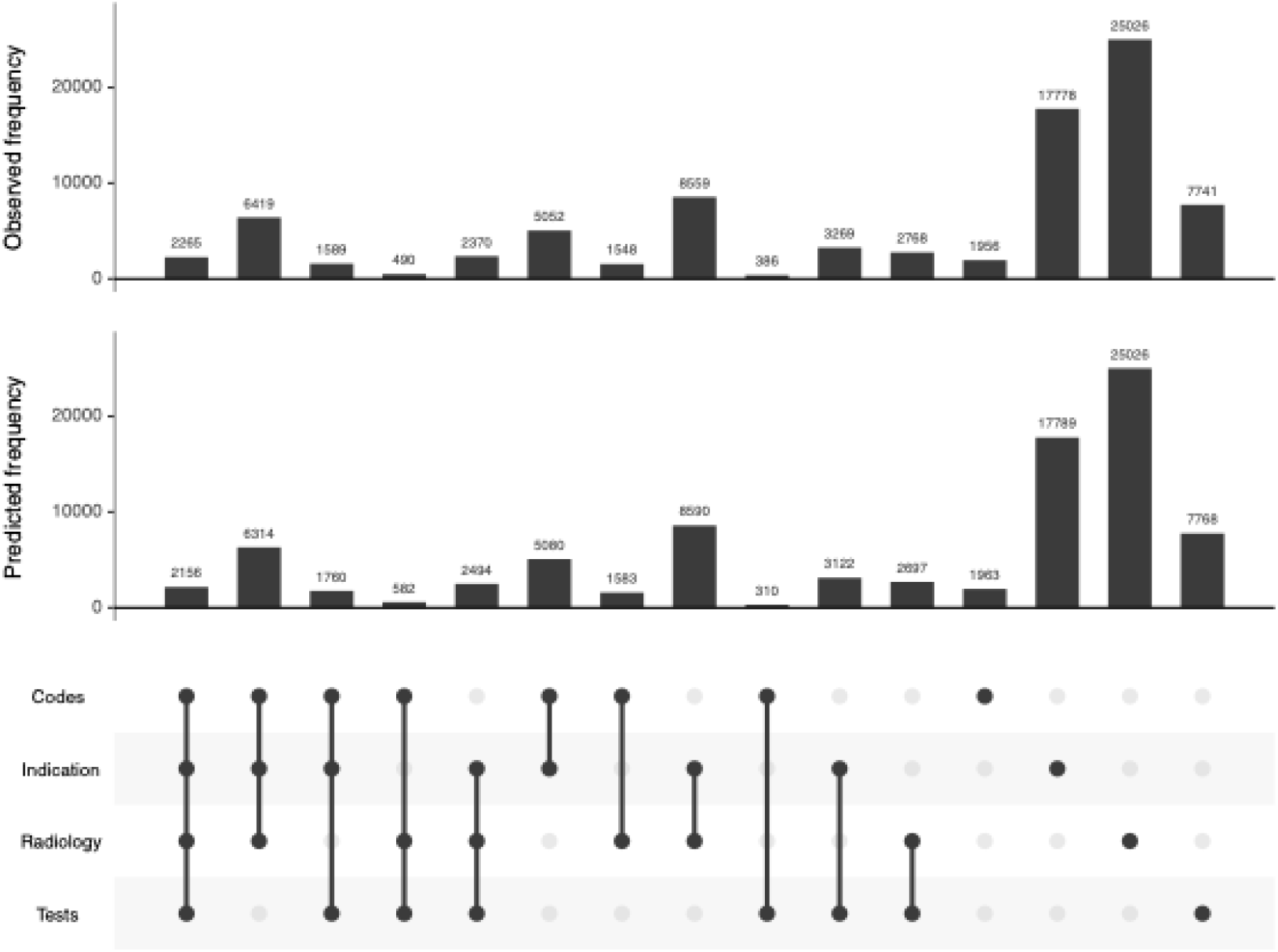
Observed and predicted frequency of each algorithm combination of community-acquired pneumonia (CAP) in Model-3 in the primary analysis. Codes: CAP primary diagnostic codes; Indication: CAP antibiotic indication; Radiology: chest X-ray report or CT scan report; Tests: shortness of breath and elevated C-reactive protein levels.

Using the alternative less informative priors, results remained stable, indicating that the choice of priors had little impact on the results (**Table S5)**. Sensitivity analyses of alternative approaches to missing data showed similar results (**Supplementary Results, Table S6-8**). We also considered a higher oxygen saturation threshold (<95%) in tests, which improved the estimated sensitivity to 0.374 in the primary analysis, although the specificity slightly decreased to 0.963.

The proportion of admissions with CAP diagnostic codes was approximately 5% from 2016 to 2019, dropping to ∼3% in 2020-2021, before increasing again in 2022 and 2023. The estimated CAP prevalence (latent class) followed a similar trend, but with higher absolute values, ranging from 13-14% between 2016 and 2019, declining to 11% in 2020, and rising to 12-13% in 2022 and 2023 (**Figure 4**).

**Figure 4.**
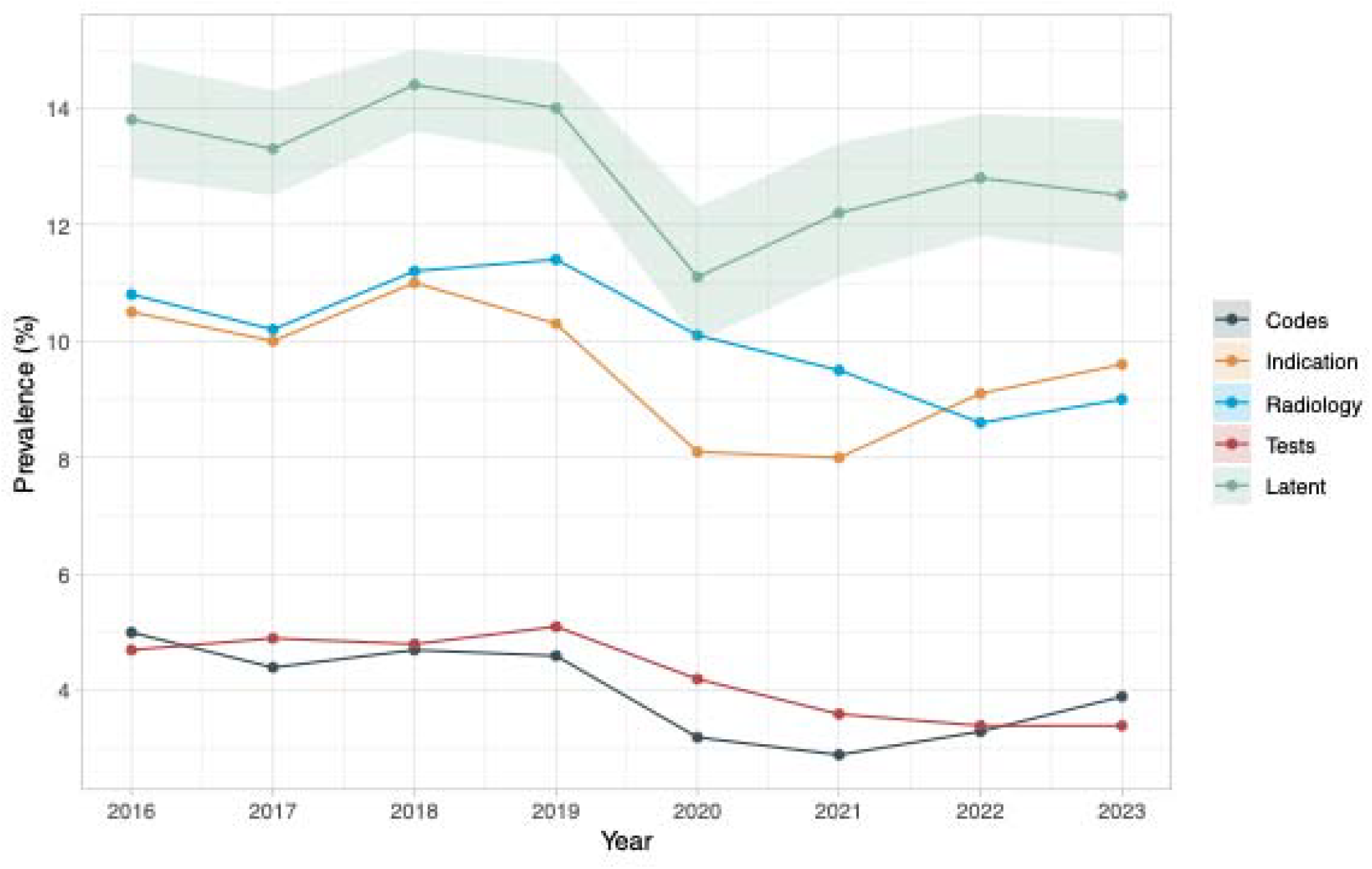
Proportion of positive results in each algorithm and latent prevalences by year. Codes: CAP primary diagnostic codes; Indication: CAP antibiotic indication; Radiology: chest X-ray report or CT scan report; Tests: shortness of breath and elevated C-reactive protein levels; Latent: latent prevalence estimated from the model. Shaded area shows the 95% credible intervals for the latent prevalence estimated from the model.

## Discussion

Identifying CAP patients retrospectively for epidemiological and surveillance studies is typically based on discharge diagnostic codes, e.g. ICD-10 codes. However, using diagnostic codes risks subjectivity, variability, and inaccuracy^28^, and requires careful examination and validation before use for epidemiological research. In the absence of a gold diagnostic standard for CAP, we described a Bayesian latent class model to estimate patients’ true disease status using EHR data. This model has been previously used to identify clinical outcomes from EHRs such as traumatic brain injury^29^, osteoarthritis^30^, sepsis^25^, systemic autoimmune rheumatic diseases^31^, and more recently examine the diagnostic accuracy of SARS-CoV-2 tests^32,33^. However, its use for CAP identification has been limited.

We proposed four EHR-based algorithms for CAP identification and incorporated potential pairwise correlations between the algorithms on top of a conditional independence model. Our analysis estimated CAP prevalence as the reason for emergency admission to hospital in the studied region at 12-13%, substantially higher than estimates derived solely from discharge diagnostic codes (4-5%).

We found that the sensitivity of diagnostic codes for identifying CAP from administrative data was low, consistent with prior literature on CAP coding accuracy. For example, a Thai study examined the validity of ICD-10 primary diagnostic codes through medical record review and estimated the sensitivities for CAP codes J15.9/J18.1/J18.9 at only 37.9%/7.6%/18.3% respectively^11^. An US cross-sectional study reported that ICD-9 codes had low sensitivities (14.0%-95.9% by pathogen) to identify organism-specific pneumonia, although the specificity was high^34^. Our results add to this evidence and suggest that relying solely on primary diagnostic codes in epidemiological studies could lead to significant underestimation of CAP cases, potentially biasing results and underestimating the population-wide impact of CAP. However, the high specificity of diagnostic codes showed that almost all patients with a primary diagnostic code of CAP did have the disease. It is not immediately clear why multiple patients in our dataset had radiological evidence of CAP and were considered by the model to have CAP, but did not receive a diagnostic code for CAP. In part this may represent local coding practice requiring a diagnosis to be stated in a specific or definitive way for a code to be recorded.

Indication information from antibiotic prescriptions had a higher sensitivity than diagnostic codes alone, suggesting that free-text antibiotic indications could be used for disease identification, although the higher sensitivity was a trade off with a slightly lower specificity. Of note, the ‘indication’ written by clinicians may vary and not be limited to ‘pneumonia’ or ‘CAP’ and miss-spelling could occur. To account for this, we used a transformer-based screen followed by fuzzy string-matching multiple CAP-related keywords, providing broad coverage. Our results also align with a recent study reporting a sensitivity of 64% and specificity of 96% for antibiotic indication text for CAP^35^. The lower sensitivity from radiology may be partly because not all CAP patients were sent for a scan and some of those who had scans did not receive a formal radiologist report; these were assumed normal in primary analyses. In sensitivity analysis only including admissions with scans, sensitivity increased but specificity decreased. Radiology specificity was lower than other algorithms, suggesting our analysis of radiology reports was more likely to include false positives. One possible reason was that the negation detection method was not able to identify all negation patterns, and mistakenly classified some reports containing the CAP keywords but negated as positive.

The combination of shortness of breath and elevated CRP levels had the lowest sensitivity (0.24) among the four algorithms (although not substantially different to diagnostic codes (0.28)), even though shortness of breath is a common CAP symptom, and CRP has moderate diagnostic value^36,37^. Despite this, specificity remained high (0.988). Our algorithm was based on exemplar thresholds for respiratory rate, oxygen saturation and CRP values, rather than trying to optimize sensitivity. However, at least for respiratory rate and CRP, thresholds were relatively low. Using higher oxygen saturation thresholds (<95%) improved sensitivity, although with slightly lower specificity. In sensitivity analyses with missing data imputed or excluded, sensitivity was slightly higher (0.33-0.36), indicating that the absence of vital signs or lab measurements was not always due to patients having normal status, and could for example have reflected data quality issues rather than these never having been recorded initially.

Exploiting various information from EHR data is helpful for disease prediction and classification. Previous studies have shown that models incorporating additional administrative information—such as procedure codes, age, and length of stay—can achieve higher sensitivity and PPV for identifying CAP than using diagnostic codes alone^38^. In one previous study, coupling a text classifier based on the CXR imaging report with diagnostic codes using ‘OR’ rule increased the PPV by 20-70% while retaining a relatively high sensitivity compared with using structured EHR data alone^9^. Similarly we found that considering any of our four algorithms together substantially increased sensitivity to 0.845 whilst only reducing specificity to 0.925. Other studies using machine learning (ML) models that integrate vital signs, lung sounds, and CRP biomarkers have achieved high sensitivity (98.2%) and specificity (97.6%) for childhood CAP detection^39^. However, building predictive models usually requires a true disease label, which is difficult to obtain from administrative data without manual validation from clinicians, which is itself intrinsically challenging for CAP.

Our year-by-year analysis using BLCM found that non-COVID CAP prevalence as the reason for emergency admission to hospital remained stable at 13-14% from 2016 to 2019, with a drop in 2020, likely due to a reduction in other respiratory illness during the COVID-19 pandemic (and the competing risk of SARS-CoV-2 pneumonia, which was excluded), followed by a rise in 2022 and 2023, but still to slightly lower levels than pre-COVID. We previously reported an increasing burden of CAP hospitalisation from 1998 to 2014 in the same region based on diagnostic codes^4^; however further increases from 2016 to 2023 were not apparent in this study.

Given all four algorithms were sub-optimal on their own, we estimated the performance of outcome combinations using an ‘AND’ or ‘OR’ logical operand. Under the ‘AND’ rule, all the two, three, four outcome combinations resulted in very low sensitivity, reduced NPV, but high specificity, and PPV. On the other hand, using the ‘OR’ rule, some outcome combinations achieved higher sensitivity without impacting much on specificity. For example, if we identified CAP patients based on any positive result among the four algorithms, sensitivity increased to 0.85 while specificity remained high at 0.93.

Our study had several limitations. Microbiology data were not used, as most CAP patients never have a positive microbiological test identifying the causative organism. The keywords in the string-matching process included “LRTI” which could also capture other respiratory infections such as bronchitis or influenza. The radiology report analysis was based on keyword matching and negation detection, whose accuracy was only assessed through the BLCM. Although machine learning and natural language processing methods have been widely applied to extract CAP diagnoses from free-text radiology reports^40–42^, we did not use these methods due to the absence of labeled training data (which is challenging for CAP as described in the Introduction, and very time consuming given the size of the dataset) and the complexity of training these models. Nevertheless, BLCM can adjust for imperfect accuracies from each individual test to estimate the true disease prevalence. Also, this study only included pairwise conditional dependency, but more complex relationships among more than two algorithms might exist. However, adding more covariances to the model leads to convergence problems due to parameter identifiability issues^43^. We were not able to validate the model on an independent test dataset, therefore it is possible that algorithm definitions may not generalised to other hospitals.

In conclusion, relying on diagnostic codes alone is insufficient for accurate CAP identification and can bias epidemiological studies and lead to under-identification of CAP when using EHR data. BLCM provide a robust approach for identifying CAP from EHRs in the absence of a gold diagnostic standard. Admissions with positive results from any of the diagnostic codes, antibiotic indications, radiology reports, or clinical test data are likely to represent true CAP cases sufficiently for surveillance. Our results support that routinely collected data from EHRs can be used in conjunction with diagnostic codes to improve CAP incidence and outcome surveillance, providing a more reliable basis for public health interventions.

## Data Availability

The datasets analysed during the current study are not publicly available as they contain personal data but are available from the Infections in Oxfordshire Research Database (https://oxfordbrc.nihr.ac.uk/ research-themes-overview/antimicrobial- resistance-and-modernising-microbiology/ infections-in-oxfordshire-research-database-iord/), subject to an application and research proposal meeting the ethical and governance requirements of the Database.

## Author Contributions

The study was designed and planned by DWE and ASW. The specific analysis was designed by JW, DWE, and ASW. JW, KY, AL contributed to the statistical analysis of the data. JW, DWE, and ASW drafted the manuscript and all authors contributed to interpretation of the data and results and revised the manuscript. All authors approved the final version of the manuscript.

## Funding

This study was funded by the National Institute for Health Research (NIHR) Health Protection Research Unit in Healthcare Associated Infections and Antimicrobial Resistance at Oxford University in partnership with the UK Health Security Agency (UKHSA) (NIHR200915) and the NIHR Biomedical Research Centre, Oxford. DWE is supported by a Robertson Fellowship. The views expressed in this publication are those of the authors and not necessarily those of the NHS, the National Institute for Health Research, the Department of Health or the UKHSA.

## Acknowledgements

This work uses data provided by patients and collected by the UK’s National Health Service as part of their care and support. We thank all the people of Oxfordshire who contribute to the Infections in Oxfordshire Research Database. Research Database Team: L Butcher, H Boseley, C Crichton, DW Crook, D Eyre, O Freeman, J Gearing (community), R Harrington, K Jeffery, M Landray, A Pal, TEA Peto, TP Quan, J Robinson (community), J Sellors, B Shine, AS Walker, D Waller. Patient and Public Panel: G Blower, C Mancey, P McLoughlin, B Nichols.

## Data sharing

The datasets analysed during the current study are not publicly available as they contain personal data but are available from the Infections in Oxfordshire Research Database (https://oxfordbrc.nihr.ac.uk/research-themes-overview/antimicrobial-resistance-and-modernising-microbiology/infections-in-oxfordshire-research-database-iord/), subject to an application and research proposal meeting the ethical and governance requirements of the Database. For further details on how to apply for access to the data and for a research proposal template please.

## Declaration of interests

No author has a conflict of interest to declare.

## Notes

### Competing Interest Statement

The authors have declared no competing interest.

### Author Declarations

Research Ethics Committee, Health Research Authority and Confidentiality Advisory Group approvals (19/SC/0403,19/CAG/0144)

