## supplementary file for "Community-acquired pneumonia identification from electronic health records in the absence of a gold standard: a Bayesian latent class analysis"

### Supplementary methods

**Free text radiology reports**

The labelling rules were: 1) A report was considered ‘positive’ if it contained any of keywords indicating a possibility of CAP ('pneumonia', 'consolidation', 'infiltrate', 'airspace', 'bronchopneumonia', 'infection', 'infective', 'air bronchogram', 'density', 'pneumonic', 'abscess', 'aspiration', 'cavity'), and the keyword was not negated (negation detection was conducted using negspaCy package in Python, based on the NegEx algorithm for identifying negated findings in medical records^1^). 2) A report was considered ‘positive’ if it contained ‘weeks’, given serial imaging is often suggested to ensure a pneumonia resolves, e.g., ‘A repeat CXR in 6 weeks is suggested’. 3) A report was considered ‘negative’ if it contained ‘clear’ or ‘normal’. 4) A report was considered negative if it contained keywords indicating diseases other than CAP ('heart failure', 'oedema', 'bronchitis', 'tumour', 'cancer', 'asthma', 'fracture') and they keyword was not negated. Given these labels may be correlated or imperfect, a sophisticated label model, “Snorkel”, was used to combine the outputs of the four rules and produce a final noise-aware probabilistic label for analysis, representing the probability of CAP or its absence^2^. The label model learns weights for the labelling functions using the label matrix as input. **Table S2** shows coverage, overlaps, and conflicts of each label.

**Priors**

We assumed a uniform (0.02, 0.15) prior for the probability of having pneumonia in EHRs based on existing literature^3–5^. Priors for sensitivity and specificity were specified using beta distributions targeting specific modes and lower limits on performance based on existing literature. For Algorithm-1, we chose a mode of 0.6/lower limit of 0.35 for sensitivity, and a mode of 0.9/lower limit of 0.7 for specificity as the prior, based on existing studies on CAP coding accuracy^3,6–9^. For Algorithm-2, since information on the sensitivity/specificity of antibiotic indication was limited, we assumed a higher sensitivity and a lower specificity than diagnostic codes with wider bounds (**Table S3**). For Algorithm-3, existing literature on diagnosing CAP through radiology reports had relatively high sensitivity and specificity around 0.9^10,11^. However, these estimates were not directly applicable because of the different methods. We therefore assumed our algorithm had a good specificity but lower sensitivity, using a mode of 0.6 for sensitivity and 0.9 for specificity, and decreasing the lower bounds for both. For Algorithm-4, we used the same prior as Algorithm-1. We additionally selected two sets of less informative priors either with wider bounds in beta distributions or using uniform distributions for sensitivity analyses to examine the robustness of the posterior estimates (**Table S3**).

**Computation**

The BLCM was estimated in Just Another Gibbs Sampler (JAGS) version 4.3.0^12^ through the R interface “jagsUI”. JAGS is an implementation of a Markov Chain Monte Carlo (MCMC) algorithm called Gibbs sampling to sample the posterior distribution of a Bayesian model.

Three chains of 100,00 iterations were run. The first 10,00 iterations were discarded as the burn-in period to get to the stationary state. The chains were then thinned by taking every tenth sample to reduce autocorrelation. Convergence was checked by verifying the Gelman-Rubin statistic (≤1.1) and visual inspection of the MCMC trace plots, created using the ‘MCMCvis’ library^13^.

95% credible intervals (Crl) were calculated from the highest posterior density intervals, and means of the posterior distributions as point estimates.

### Supplementary results

In sensitivity analyses using MICE to impute missing vital signs and CRP measurements, 38,084 (7.7%) admissions had shortness of breath and elevated CRP levels at admission. The estimated CAP prevalence was 13.8% (95% Crl 13.5-14.1%), with sensitivity and specificity similar to the primary analysis (**Table S6**). In sensitivity analyses excluding admissions with missing vital signs and CRP measurements, the estimated prevalence was 12.6% (95% Crl 12.2-13.0%), with consistent sensitivity, specificity, PPV, and NPV to the main analysis (**Table S7**). In sensitivity analyses excluding admissions with missing radiology scans, the estimated prevalence increased to 15.0% (95% Crl 15.0-15.0%), with higher sensitivity and lower specificity for antibiotic indication and radiology approaches compared to primary analyses (**Table S8**).

### Supplementary Figures and Tables

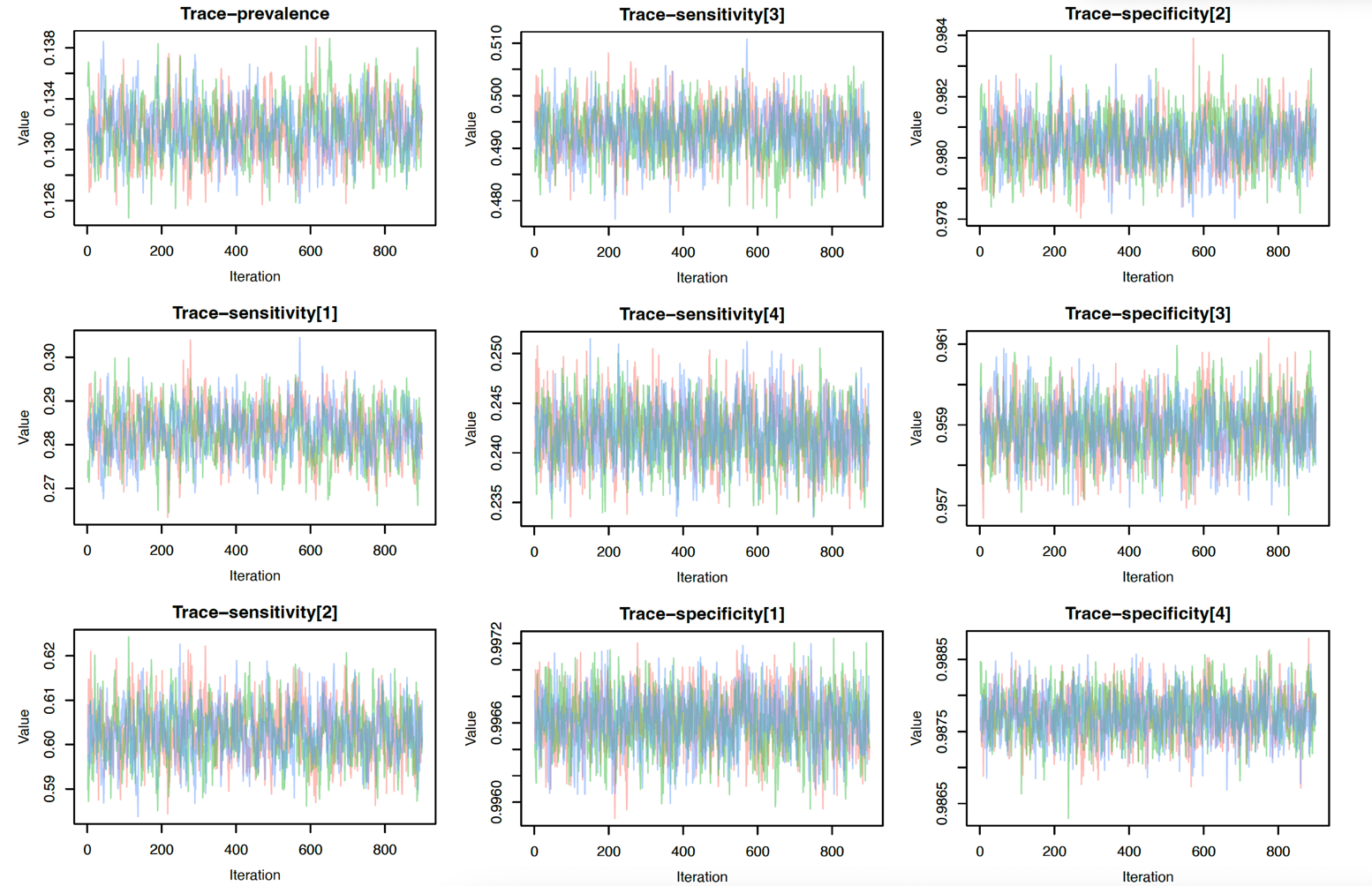

**Figure S1. Trace plots of estimated prevalence, sensitivity, and specificity of each algorithm of Model-3 in the primary analysis.** Algorithm [1] is CAP primary diagnostic codes; Algorithm [2] is CAP antibiotic indication; Algorithm [3] is chest X-ray report or CT scan report; Algorithm [4] is shortness of breath and elevated C-reactive protein levels. Trace plots indicate good convergence.

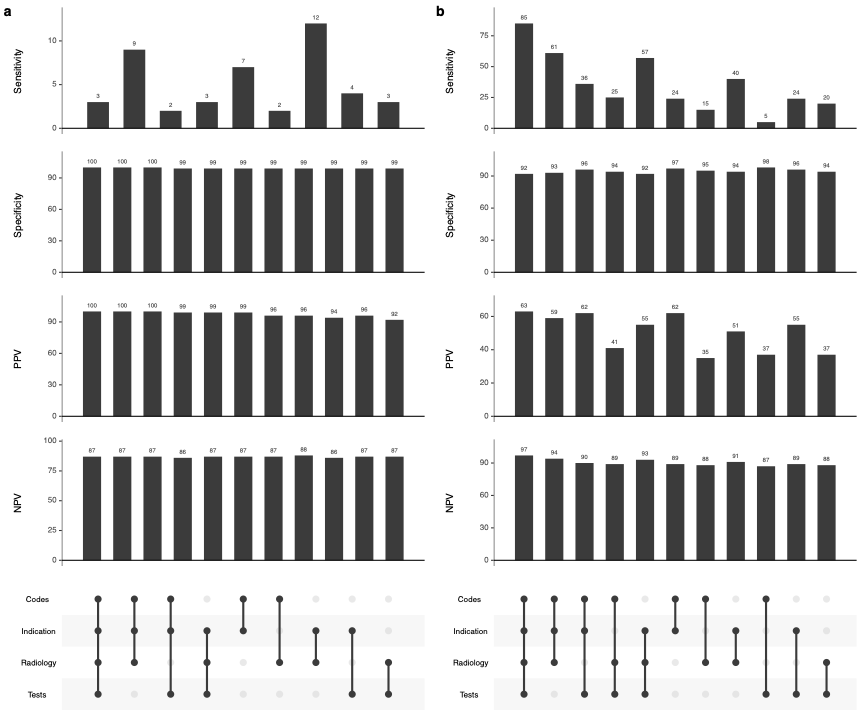

**Figure S2. Posterior predicted sensitivity, specificity, PPV, NPV for each algorithm combination under the ‘AND’ rule (a) and ‘OR’ rule (b) in Model-3 in the primary analysis.** PPV: positive predictive value; NPV: negative predictive value. Numbers are shown in **Table 2**.

| ICD10 code | ICD10 diagnosis | Frequency | Proportion (%) |
| --- | --- | --- | --- |
| J189 | Pneumonia, unspecified organism | 9482 | 48.12 |
| J181 | Lobar pneumonia, unspecified organism | 9412 | 47.76 |
| J180 | Bronchopneumonia, unspecified organism | 280 | 1.42 |
| J159 | Bacterial pneumonia, unspecified | 144 | 0.73 |
| J13 | Pneumonia due to Streptococcus pneumoniae | 94 | 0.48 |
| J151 | Pneumonia due to Pseudomonas | 64 | 0.32 |
| J154 | Pneumonia due to other streptococci | 48 | 0.24 |
| J851 | Abscess of lung with pneumonia | 37 | 0.19 |
| J14 | Pneumonia due to Haemophilus influenzae | 36 | 0.18 |
| J152 | Pneumonia due to staphylococcus | 20 | 0.10 |
| A481 | Legionnaires disease | 19 | 0.10 |
| J150 | Pneumonia due to klebsiella pneumoniae | 15 | 0.08 |
| J188 | Other pneumonia, unspecified organism | 13 | 0.07 |
| J157 | Pneumonia due to Mycoplasma pneumoniae | 11 | 0.06 |
| J155 | Pneumonia due to Escherichia coli | 9 | 0.05 |
| J156 | Pneumonia due to other Gram-negative bacteria | 6 | 0.03 |
| J850 | Gangrene and necrosis of lung | 6 | 0.03 |
| J158 | Pneumonia due to other specified bacteria | 4 | 0.02 |
| J153 | Pneumonia due to streptococcus, group B | 3 | 0.02 |
| J168 | Pneumonia due to other specified infectious organisms | 2 | 0.01 |

**Table S1. Frequency and proportion of ICD-10 diagnostic codes for CAP in 19,705 admissions with primary diagnosis of CAP.** CAP: community-acquired pneumonia.

| **Algorithm** | **Labelling rules** | **Polarity** | **Coverage** | **Overlaps** | **Conflicts** |
| --- | --- | --- | --- | --- | --- |
| 1 | Positive keywords* | 1 | 0.208 | 0.083 | 0.038 |
| 2 | Clear | 0 | 0.376 | 0.288 | 0.002 |
| 3 | Normal | 0 | 0.379 | 0.302 | 0.002 |
| 4.1 | Week | 1 | 0.056 | 0.056 | 0.008 |
| 4.2 | Weeks | 1 | 0.054 | 0.054 | 0.008 |
| 5 | Negative keywords** | 0 | 0.132 | 0.093 | 0.039 |

**Table S2. Coverage, overlaps, and conflicts of the labelling rules used for identifying community-acquired pneumonia from free text chest X-ray or CT reports.** *Positive keywords: 'pneumonia', 'consolidation', 'infiltrate', 'airspace', 'bronchopneumonia', 'infection', 'infective', 'air bronchogram', 'density', 'pneumonic', 'abscess', 'aspiration', 'cavity'; **Negative keywords: 'heart failure', 'oedema', 'bronchitis', 'tumour', 'cancer', 'asthma', 'fracture'.

| **Algorithm** | **Parameter** | **Primary prior set** | **Alternative prior set 1** | **Alternative prior set 2** |
| --- | --- | --- | --- | --- |
|  | CAP prevalence | Uniform (0.02, 0.15) | Uniform (0.02, 0.15) | Uniform (0.02, 0.15) |
| CAP primary diagnostic codes | Sensitivity | Mode=0.60, 95% sure >0.35; Beta(7.01, 5.01) | Mode=0.50, 95% sure >0.20; Beta(3.26, 3.26) | Uniform (0.2, 0.8) |
|  | Specificity | Mode=0.90, 95% sure >0.70; Beta(15.03, 2.56) | Mode=0.90, 95% sure >0.50; Beta(5.38, 1.49) | Uniform (0.6,1) |
| Antibiotics Indication | Sensitivity | Mode=0.80, 95% sure >0.50; Beta(7.55, 2.64) | Mode=0.70, 95% sure >0.20; Beta(2.26, 1.54) | Uniform (0.2, 0.8) |
|  | Specificity | Mode=0.85, 95% sure >0.50; Beta(6.25, 1.93) | Mode=0.90, 95% sure >0.50; Beta(5.38, 1.49) | Uniform (0.6,1) |
| Radiology report | Sensitivity | Mode=0.60, 95% sure >0.35; Beta(7.01, 5.01) | Mode=0.50, 95% sure >0.20; Beta(3.26, 3.26) | Uniform (0.2, 0.8) |
|  | Specificity | Mode=0.90, 95% sure >0.60; Beta(8.30, 1.81) | Mode=0.90, 95% sure >0.50; Beta(5.38, 1.49) | Uniform (0.6,1) |
| Test results | Sensitivity | Mode=0.60, 95% sure >0.35; Beta(7.01, 5.01) | Mode=0.50, 95% sure >0.20; Beta(3.26, 3.26) | Uniform (0.2, 0.8) |
|  | Specificity | Mode=0.90, 95% sure >0.70; Beta(15.03, 2.56) | Mode=0.90, 95% sure >0.50; Beta(5.38, 1.49) | Uniform (0.6,1) |

**Table S3. Priors for the prevalence of community-acquired pneumonia, sensitivities and specificities of the four algorithms in the Bayesian latent class model.**

| **Algorithms** | | | | | **Observed frequency** | | **Predicted frequency** | | | |
| --- | --- | --- | --- | --- | --- | --- | --- | --- | --- | --- |
| **Primary codes** | **Antibiotic indication** | **Radiology report** | **Test results** |  | | **Model1** | | **Model2** | **Model3** | **Model4** |
| 1 | 1 | 1 | 0 | 6419 | | 5657 | | 6347 | 6314 | 6298 |
| 1 | 1 | 0 | 1 | 2265 | | 2084 | | 2108 | 2156 | 2167 |
| 1 | 1 | 1 | 1 | 1589 | | 1809 | | 1704 | 1760 | 1749 |
| 1 | 0 | 1 | 1 | 490 | | 716 | | 529 | 582 | 586 |
| 0 | 1 | 1 | 1 | 2370 | | 3128 | | 2671 | 2494 | 2500 |
| 1 | 1 | 0 | 0 | 5052 | | 4919 | | 5150 | 5080 | 5096 |
| 0 | 1 | 0 | 1 | 3269 | | 2893 | | 3105 | 3122 | 3119 |
| 1 | 0 | 1 | 0 | 1548 | | 1957 | | 1641 | 1583 | 1580 |
| 1 | 0 | 0 | 1 | 8559 | | 9042 | | 8415 | 8590 | 8593 |
| 0 | 1 | 1 | 0 | 386 | | 626 | | 303 | 310 | 308 |
| 0 | 0 | 1 | 1 | 2768 | | 1475 | | 2693 | 2697 | 2691 |
| 0 | 1 | 0 | 0 | 17778 | | 17763 | | 17806 | 17789 | 17789 |
| 0 | 0 | 0 | 1 | 7741 | | 8165 | | 7770 | 7768 | 7770 |
| 1 | 0 | 0 | 0 | 25026 | | 25385 | | 25036 | 25026 | 25036 |
| 0 | 0 | 1 | 0 | 1956 | | 1936 | | 1961 | 1963 | 1963 |
| 0 | 0 | 0 | 0 | 404465 | | 404123 | | 404441 | 404447 | 404436 |

**Table S4. Cross-classified results for the observed frequency and the predicted frequency of each algorithm combination under four models in the primary analysis.**

| **Parameter** | **Alternative prior 1** | **Alternative prior 2** |
| --- | --- | --- |
| prevalence | 0.132 (0.128-0.136) | 0.132 (0.128-0.136) |
| **Primary codes** | | |
| sensitivity | 0.282 (0.272-0.292) | 0.282 (0.271-0.292) |
| specificity | 0.997 (0.996-0.997) | 0.997 (0.996-0.997) |
| PPV | 0.926 (0.917-0.935) | 0.926 (0.917-0.935) |
| NPV | 0.901 (0.897-0.905) | 0.901 (0.896-0.905) |
| **Antibiotic indication** | | |
| sensitivity | 0.602 (0.590-0.613) | 0.601 (0.589-0.613) |
| specificity | 0.981 (0.979-0.982) | 0.981 (0.979-0.982) |
| PPV | 0.825 (0.812-0.839) | 0.826 (0.811-0.840) |
| NPV | 0.942 (0.939-0.945) | 0.942 (0.938-0.945) |
| **Radiology report** | | |
| sensitivity | 0.492 (0.483-0.501) | 0.492 (0.483-0.501) |
| specificity | 0.959 (0.958-0.960) | 0.959 (0.958-0.960) |
| PPV | 0.646 (0.634-0.658) | 0.646 (0.634-0.659) |
| NPV | 0.925 (0.922-0.929) | 0.925 (0.922-0.929) |
| **Test results** | | |
| sensitivity | 0.241 (0.236-0.247) | 0.241 (0.236-0.247) |
| specificity | 0.988 (0.987-0.988) | 0.988 (0.987-0.988) |
| PPV | 0.750 (0.738-0.763) | 0.751 (0.737-0.764) |
| NPV | 0.895 (0.892-0.899) | 0.895 (0.891-0.899) |
| **Covariance** | | |
| cov12\|D=1 | 0.066 (0.063-0.069) | 0.066 (0.063-0.069) |
| cov13\|D=1 | 0.024 (0.021-0.027) | 0.024 (0.021-0.027) |
| cov14\|D=1 | 0.006 (0.004-0.008) | 0.006 (0.004-0.008) |
| **Model comparison** | | |
| DIC | 245 | 246 |

**Table S5. Posterior predicted results under Model 3 in the primary analysis using alternative prior sets.** Alternative prior information is summarised in Table S3.

| **Parameter** | **Model 1** | **Model 2** | **Model 3** | **Model 4** |
| --- | --- | --- | --- | --- |
| prevalence | 0.107 (0.106-0.109) | 0.132 (0.130-0.135) | 0.138 (0.135-0.141) | 0.137 (0.134-0.141) |
| **Primary codes** | | | | |
| sensitivity | 0.372 (0.366-0.377) | 0.289 (0.283-0.296) | 0.275 (0.267-0.283) | 0.275 (0.267-0.282) |
| specificity | 1.000 (0.999-1.000) | 0.998 (0.998-0.998) | 0.997 (0.997-0.998) | 0.997 (0.997-0.997) |
| PPV | 0.992 (0.988-0.997) | 0.953 (0.948-0.958) | 0.942 (0.935-0.948) | 0.938 (0.928-0.946) |
| NPV | 0.930 (0.928-0.931) | 0.902 (0.899-0.904) | 0.896 (0.893-0.899) | 0.896 (0.893-0.900) |
| **Antibiotic indication** | | | | |
| sensitivity | 0.785 (0.779-0.792) | 0.663 (0.654-0.671) | 0.653 (0.643-0.662) | 0.653 (0.644-0.663) |
| specificity | 0.968 (0.967-0.969) | 0.971 (0.970-0.973) | 0.974 (0.972-0.975) | 0.974 (0.972-0.975) |
| PPV | 0.747 (0.740-0.754) | 0.780 (0.771-0.788) | 0.799 (0.787-0.810) | 0.797 (0.786-0.809) |
| NPV | 0.974 (0.973-0.975) | 0.950 (0.948-0.952) | 0.946 (0.944-0.949) | 0.946 (0.944-0.949) |
| **Radiology report** | | | | |
| sensitivity | 0.520 (0.514-0.526) | 0.493 (0.486-0.499) | 0.483 (0.475-0.490) | 0.483 (0.476-0.491) |
| specificity | 0.950 (0.949-0.951) | 0.959 (0.958-0.960) | 0.960 (0.959-0.962) | 0.960 (0.959-0.961) |
| PPV | 0.554 (0.548-0.560) | 0.649 (0.640-0.657) | 0.661 (0.651-0.671) | 0.660 (0.651-0.670) |
| NPV | 0.943 (0.942-0.944) | 0.925 (0.923-0.927) | 0.921 (0.918-0.923) | 0.921 (0.918-0.924) |
| **Test results** | | | | |
| sensitivity | 0.383 (0.378-0.389) | 0.365 (0.360-0.370) | 0.356 (0.350-0.362) | 0.356 (0.350-0.363) |
| specificity | 0.959 (0.958-0.960) | 0.966 (0.966-0.967) | 0.967 (0.966-0.968) | 0.967 (0.966-0.968) |
| PPV | 0.530 (0.523-0.536) | 0.624 (0.615-0.633) | 0.633 (0.623-0.642) | 0.632 (0.623-0.641) |
| NPV | 0.928 (0.927-0.930) | 0.909 (0.907-0.911) | 0.904 (0.901-0.907) | 0.904 (0.901-0.907) |
| **Covariance** | | | | |
| cov12\|D=1 | NA | 0.058 (0.056-0.060) | 0.061 (0.058-0.063) | 0.061 (0.058-0.063) |
| cov13\|D=1 | NA | 0.018 (0.016-0.020) | 0.022 (0.020-0.024) | 0.022 (0.020-0.025) |
| cov14\|D=1 | NA | NA | 0.007 (0.005-0.009) | 0.007 (0.005-0.009) |
| cov12\|D=0 | NA | NA | NA | 0.000 (0.000-0.000) |
| **Model comparison** | | | | |
| DIC | 2356 | 417 | 384 | 388 |

**Table S6. Posterior predicted sensitivity, specificity, PPV, NPV, covariance, and model comparison among four models with different covariance structures in the sensitivity analysis with missing vitals and CRP measurements imputed.** PPV: positive predictive value; NPV: negative predictive value; DIC: deviance information criteria.

| **Parameter** | **Model 1** | **Model 2** | **Model 3** | **Model 4** |
| --- | --- | --- | --- | --- |
| prevalence | 0.097 (0.095-0.098) | 0.121 (0.118-0.123) | 0.126 (0.122-0.130) | 0.126 (0.122-0.129) |
| **Primary codes** | | | | |
| sensitivity | 0.388 (0.381-0.395) | 0.299 (0.291-0.307) | 0.282 (0.272-0.292) | 0.280 (0.270-0.291) |
| specificity | 1.000 (1.000-1.000) | 0.998 (0.998-0.998) | 0.997 (0.997-0.998) | 0.997 (0.996-0.998) |
| PPV | 0.995 (0.991-0.999) | 0.954 (0.948-0.961) | 0.940 (0.932-0.949) | 0.932 (0.914-0.945) |
| NPV | 0.938 (0.937-0.940) | 0.912 (0.909-0.915) | 0.906 (0.902-0.910) | 0.906 (0.902-0.910) |
| **Antibiotic indication** | | | | |
| sensitivity | 0.733 (0.725-0.740) | 0.612 (0.602-0.623) | 0.601 (0.589-0.612) | 0.601 (0.590-0.613) |
| specificity | 0.976 (0.975-0.977) | 0.979 (0.977-0.980) | 0.981 (0.979-0.982) | 0.980 (0.979-0.982) |
| PPV | 0.765 (0.757-0.774) | 0.796 (0.786-0.806) | 0.817 (0.804-0.831) | 0.814 (0.800-0.829) |
| NPV | 0.971 (0.970-0.973) | 0.948 (0.946-0.951) | 0.944 (0.942-0.947) | 0.945 (0.942-0.948) |
| **Radiology report** | | | | |
| sensitivity | 0.532 (0.525-0.539) | 0.500 (0.492-0.508) | 0.490 (0.481-0.499) | 0.491 (0.481-0.501) |
| specificity | 0.948 (0.948-0.949) | 0.957 (0.956-0.958) | 0.958 (0.957-0.960) | 0.958 (0.957-0.960) |
| PPV | 0.525 (0.518-0.532) | 0.615 (0.605-0.625) | 0.629 (0.618-0.641) | 0.629 (0.617-0.640) |
| NPV | 0.950 (0.948-0.951) | 0.933 (0.931-0.936) | 0.929 (0.926-0.932) | 0.929 (0.926-0.932) |
| **Test results** | | | | |
| sensitivity | 0.359 (0.353-0.364) | 0.337 (0.331-0.343) | 0.327 (0.320-0.334) | 0.328 (0.320-0.335) |
| specificity | 0.978 (0.977-0.978) | 0.984 (0.983-0.985) | 0.984 (0.984-0.985) | 0.984 (0.984-0.985) |
| PPV | 0.633 (0.624-0.642) | 0.741 (0.729-0.753) | 0.751 (0.738-0.764) | 0.750 (0.738-0.763) |
| NPV | 0.934 (0.933-0.936) | 0.915 (0.913-0.918) | 0.910 (0.907-0.914) | 0.911 (0.907-0.914) |
| **Covariance** | | | | |
| cov12\|D=1 | NA | 0.063 (0.060-0.065) | 0.066 (0.063-0.068) | 0.065 (0.062-0.068) |
| cov13\|D=1 | NA | 0.019 (0.016-0.021) | 0.023 (0.020-0.026) | 0.024 (0.021-0.027) |
| cov14\|D=1 | NA | NA | 0.008 (0.005-0.010) | 0.008 (0.005-0.011) |
| cov12\|D=0 | NA | NA | NA | 0.000 (0.000-0.001) |
| **Model comparison** | | | | |
| DIC | 1700 | 269 | 241 | 243 |

**Table S7. Posterior predicted sensitivity, specificity, PPV, NPV, covariance, and model comparison among four models with different covariance structures in the sensitivity analysis removing admission with missing vitals and CRP measurements.** PPV: positive predictive value; NPV: negative predictive value; DIC: deviance information criteria.

| **Parameter** | **Model 1** | **Model 2** | **Model 3** | **Model 4** |
| --- | --- | --- | --- | --- |
| prevalence | 0.150 (0.150-0.150) | 0.150 (0.150-0.150) | 0.150 (0.150-0.150) | 0.150 (0.150-0.150) |
| **Primary codes** | | | | |
| sensitivity | 0.482 (0.475-0.488) | 0.465 (0.457-0.472) | 0.465 (0.458-0.472) | 0.402 (0.393-0.410) |
| specificity | 0.997 (0.997-0.998) | 0.996 (0.995-0.997) | 0.996 (0.995-0.997) | 0.983 (0.981-0.984) |
| PPV | 0.970 (0.964-0.976) | 0.954 (0.948-0.961) | 0.954 (0.948-0.961) | 0.804 (0.790-0.819) |
| NPV | 0.916 (0.915-0.917) | 0.913 (0.912-0.914) | 0.913 (0.912-0.914) | 0.903 (0.902-0.904) |
| **Antibiotic indication** | | | | |
| sensitivity | 0.810 (0.803-0.816) | 0.777 (0.768-0.786) | 0.777 (0.768-0.786) | 0.739 (0.730-0.749) |
| specificity | 0.927 (0.925-0.928) | 0.923 (0.922-0.925) | 0.923 (0.922-0.925) | 0.914 (0.912-0.916) |
| PPV | 0.661 (0.655-0.666) | 0.642 (0.635-0.649) | 0.642 (0.635-0.649) | 0.603 (0.595-0.610) |
| NPV | 0.965 (0.964-0.966) | 0.959 (0.957-0.961) | 0.959 (0.957-0.961) | 0.952 (0.950-0.954) |
| **Radiology report** | | | | |
| sensitivity | 0.654 (0.648-0.660) | 0.664 (0.657-0.671) | 0.664 (0.657-0.672) | 0.697 (0.688-0.705) |
| specificity | 0.849 (0.847-0.851) | 0.853 (0.851-0.855) | 0.853 (0.851-0.855) | 0.857 (0.855-0.859) |
| PPV | 0.432 (0.428-0.437) | 0.443 (0.438-0.448) | 0.443 (0.438-0.448) | 0.462 (0.456-0.468) |
| NPV | 0.933 (0.932-0.934) | 0.935 (0.934-0.936) | 0.935 (0.934-0.936) | 0.941 (0.940-0.943) |
| **Test results** | | | | |
| sensitivity | 0.278 (0.272-0.283) | 0.284 (0.278-0.290) | 0.284 (0.278-0.290) | 0.304 (0.298-0.311) |
| specificity | 0.958 (0.957-0.959) | 0.960 (0.959-0.962) | 0.960 (0.959-0.961) | 0.963 (0.962-0.964) |
| PPV | 0.540 (0.531-0.548) | 0.559 (0.549-0.568) | 0.558 (0.549-0.568) | 0.594 (0.584-0.604) |
| NPV | 0.883 (0.882-0.884) | 0.884 (0.883-0.885) | 0.884 (0.883-0.885) | 0.887 (0.886-0.888) |
| **Covariance** | | | | |
| cov12\|D=1 | NA | 0.025 (0.021-0.029) | 0.025 (0.020-0.029) | 0.033 (0.029-0.037) |
| cov13\|D=1 | NA | 0.002 (0.000-0.006) | 0.003 (0.000-0.006) | 0.025 (0.021-0.029) |
| cov14\|D=1 | NA | NA | 0.000 (0.000-0.000) | 0.000 (0.000-0.001) |
| cov12\|D=0 | NA | NA | NA | 0.011 (0.010-0.012) |
| **Model comparison** | | | | |
| DIC | 1343 | 1232 | 1236 | 853 |

**Table S8.** **Posterior predicted sensitivity, specificity, PPV, NPV, covariance, and model comparison among four models with different covariance structures in the sensitivity analysis removing admissions without radiology scans.** PPV: positive predictive value; NPV: negative predictive value; DIC: deviance information criteria.
